# Inequalities in Household Access to Insecticide-Treated Nets and Indoor Residual Spraying in 34 African Countries

**DOI:** 10.1101/2025.09.23.25336506

**Authors:** Abdul-Wahab Inusah, Collins Gbeti, Temple O. Jagha, Abukari Wumbei, Shamsu-Deen Ziblim

## Abstract

**Background:** Malaria control through insecticide-treated nets (ITNs) and indoor residual spraying (IRS) is essential in sub-Saharan Africa. This study assesses wealth status and residence-based inequalities in household access to these interventions across 34 African countries.

**Methods:** A cross-sectional analytical design was used, drawing on nationally representative household surveys (Demographic and Health Surveys, Malaria Indicator Surveys, multiple Indicator Cluster Surveys) collected between 2005 and 2018. Data from the WHO Health Equity Monitor provided equity-disaggregated indicators on ITN/IRS coverage by wealth quintile and place of residence (rural/urban). The primary outcome was the percentage of households with at least one ITN and/or IRS coverage in the previous 12 months. We calculated both the percentage and disparities of inequalities across four inequality dimensions: difference (D), ratio (R), population-attributable risk (PAR), and population-attributable fraction (PAF)

**Results:** The ITN and/or IRS coverage varied widely from countries, highest in Mali (93.6%), Benin (92.0%), Burkina Faso (89.8%), and Rwanda (89.2%), and lowest in Ethiopia (3.4%) and Eswatini (4.4%). Wealth-related inequalities included pro-rich patterns with Burundi reporting the largest absolute difference (D = 33.6 percentage points), ratio (R = 2.1), population attributable risk (PAR = 17.5), and population attributable fraction (PAF = 37.5%). Other countries with notable pro-rich disparities included São Tomé and Príncipe (D = 28.8; R = 1.7; PAR = 12.1; PAF = 20.0%) and Malawi (D = 24.2; R = 1.5; PAR = 12.2; PAF = 20.8%). The strong pro-poor patterns were observed in Namibia (D = –36.5; R = 0.3), Gambia (D = –28.6; R = 0.7), and Nigeria (D = –28.2; R = 0.7), with marginal PAF, indicating minimal inequality-related coverage loss. Rural-urban disparities also varied, with Burundi showing a pro-urban absolute difference (D = 20.3), ratio (R = 1.5), PAR (18.1), and PAF (38.7%), whereas Zimbabwe (D = –31.5; R = 0.5) and Namibia (D = –32.7; R = 0.3) exhibited pro-rural advantages.

**Conclusions:** The study shows persistent wealth-related and geographical inequalities in access to ITNs and IRS across sub-Saharan Africa. Targeted efforts aimed at reducing these disparities are essential to achieving equitable and universal malaria prevention coverage consistent with global malaria and Sustainable Development Goal targets.

## Introduction

Despite enormous progress made over the last two decades in the world, malaria continues to be a leading public health challenge in Africa, particularly in sub-Saharan Africa (1). Recent estimates emphasise the disproportionate burden of the disease in Africa. For instance, the magnitude of malaria was approximately 212 million cases in the world, with roughly 90% of them coming from sub-Saharan Africa in 2015 (2). The African Region of WHO has always been the territory bearing the most significant health burden due to malaria in terms of morbidity and mortality (1,3). In 2022, approximately 94% of malaria cases in the world (233 million) and 95% of malaria deaths (about 580,000 deaths) occurred in Africa (4,5). Children under five years stand to be the most vulnerable, constituting around 70–78% of malaria deaths (2).

Efforts have been taken to reduce malaria incidence and mortality since 2000. For instance, infection rates decreased by 21% from 2010 to 2015, while deaths during the same period fell by 29% (2). However, in recent years, progress seems to have stalled. In the period since around 2015, cases and deaths, which had been falling, have now plateaued, leaving Africa lagging by more than 50% toward the achievement of the 2025 milestones of the WHO Global Technical Strategy for Malaria (2016–2030) (6). This lamentable pause, coupled with continued high case burdens in highly populated countries such as Nigeria and the Democratic Republic of Congo, places an even higher need on scaling up proven interventions and plugging the resultant gaps (7). Further gains will require high intervention coverage but will equally require coverage to be equitably distributed among all groups of populations, which is a cardinal tenet of health equity and of the “leave no one behind” pledge of the Sustainable Development Goals (SDGs) (8). The WHO has also called for maximising the public health impact of malaria prevention, emphasising universal access to effective interventions.

Insecticide-treated nets (ITNs) and indoor residual spraying (IRS), despite a plethora of newer applications and techniques, inadvertently remain the two old yet basic interventions for vector control in malaria (8,9). ITNs are mesh bed nets treated with long-lasting insecticides that repel and kill mosquitoes to protect humans from infectious bites while they sleep. IRS represents the periodic spraying of interior walls and surfaces of homes with residual insecticides, killing mosquitoes that rest indoors (2). The utility of these methods on the blocking of malaria transmission and alleviation of disease burden cannot be downplayed (10). The major contributors to a dramatic fall in malaria since 2000 have been ITNs and IRS together. A critical analysis estimated that of the 663 million malaria cases averted in sub-Saharan Africa between 2001 and 2015, about 78% were averted through ITN and IRS intervention scale-up (2,11).

Historically, global malaria strategies had revolved around achieving universal coverage of ITNs and IRS (12). The Roll Back Malaria Global Action Plan from 2008 and the WHO Global Technical Strategy for Malaria 2016–2030 chart ambitious targets for coverage, with universal access to malaria prevention presumed as granted for all populations at risk (13). WHO defines “universal coverage” for vector control as universal access to and use of effective vector control interventions (ITNs and/or IRS) by at-risk populations (14). In practical terms, goals include the distribution of enough ITNs to cover all sleeping areas and ensuring that 100% of targeted-area households are protected through IRS (9). Maintaining this level of coverage aligns with the first pillar of the global malaria strategy (15), which emphasises the shared belief that transmission decline cannot occur without high coverage with ITNs/IRS.

By the mid-2010s, billions of mosquito nets had been delivered to the malaria-endemic countries. For instance, in 2015, more than 60 million nets were distributed across Nigeria in one campaign, and dozens of countries were implementing IRS programs (16). Recent WHO records show that in 2022, about 56% of young children and pregnant women in sub-Saharan Africa were sleeping under an ITN, about the same rate as in 2015 (17,18). Even less could be said of IRS coverage: protection of the world population by IRS declined from almost 5% in 2010 to just 2% in 2019 (3,19). In recent years, some countries have, due to lack of resources, operational challenges, and insecticide resistance, been forced to scale down IRS interventions (10). They, therefore, undercut the full potential impact of these interventions due to pronounced coverage gaps in certain countries and population groups (10,20)

Such inequality in the coverage of malaria interventions has indeed become a primary concern because national-level aggregate trends can often camouflage the disparities within socioeconomic and geographic subgroups (21). Particularly noteworthy are wealth and residential location-based inequalities, meaning that disparities in coverage exist between richer and poorer households as well as urban and rural households. Such differentials have been widely documented and increasingly recognized as impediments to the achievement of control and elimination targets for malaria (22). The poorest and rural populations bear the highest malaria burdens and have diminished access to tools of prevention and services. For example, an analysis by the WHO and the Global Fund highlighted the fact that in most countries, the poorest, least-educated, and rural groups remain disadvantaged in developing HIV, TB, and malaria interventions (15).

Before 2005, during the malaria-control era, ITNs were distributed forcefully from health facilities or commercial outlets, concentrating mainly on urban and wealthier communities with access to such channels. However, the change in policies toward mass free distribution campaigns around 2008-2010 improved the equity in ITN ownership in many countries. On one hand, various sub-Saharan African countries have implemented programs to promote pro-poor coverage of ITNs, that is, the poorest households are as likely, or even more likely, to possess an ITN than the richest ones, primarily due to intentional targeting of distribution to high-burden, low-income rural areas (23). Similarly, some studies have reported a narrowing of wealth gaps in net use over time, witnessing 13 out of 25 countries achieving principally equitable ITN coverage by the dwellers’ wealth in the mid-2010s. Inequitable access to malaria prevention generates ethical and human rights questions but also presents epidemiological concerns because subgroups with low coverage act as a reservoir that sustains transmission and seeds resurgences, thus undermining the overarching objectives of control programs (23). “We must focus more on reaching the poor, rural, and least-educated populations who bear the brunt of these diseases,” Dr. Tedros Ghebreyesus, Director-General of WHO, stated (23). In the context of malaria, this means identifying and addressing the gaps in coverage among the underserved. While previous studies have considered equity trends in malaria interventions, including single-country analyses and earlier multi-country comparisons, there is a need for updated evidence that spans a wide range of countries and includes the most recent available data. This study therefore investigated wealth and residential inequalities in household access to insecticide-treated nets and indoor residual spraying in 34 African Countries.

## Materials and Methods

### Study Design and Setting

This was a cross-sectional, analytical study leveraging nationally representative household survey data from multiple countries in sub-Saharan Africa. The primary focus was to assess socioeconomic and residence-based inequalities in access to malaria prevention tools, specifically households with at least one insecticide-treated mosquito net (ITN) and/or having received indoor residual spraying (IRS) within the past 12 months. The study spanned 34 countries, with data ranging from 2005 to 2018, depending on the most recent and available nationally representative survey for each country.

### Data Sources

Data were extracted from the World Health Organization’s (WHO) Health Equity Monitor (HEM) and Global Health Observatory (GHO), which compile standardized indicators from Demographic and Health Surveys (DHS), Malaria Indicator Surveys (MIS), and Multiple Indicator Cluster Surveys (MICS). These surveys are conducted in collaboration with national statistical offices and ministries of health and employ stratified two-stage cluster sampling methods to ensure national representativeness. The data used included equity-disaggregated indicators on ITN/IRS coverage, wealth quintiles, and place of residence (rural vs. urban), along with survey year and population denominators.

### Study Population and Inclusion Criteria

The unit of analysis was the household. Only households from countries with complete information on the indicator “Households with at least one ITN and/or IRS in the past 12 months expressed as percentage” disaggregated by wealth quintile and residence were included. Countries with outdated, incomplete, or inconsistent disaggregated indicators were excluded.

### Outcome Variable

The primary outcome variable was the percentage of households with access to at least one ITN and/or that had received IRS in the past 12 months. This reflects a composite measure of household-level coverage with malaria vector control interventions and aligns with global malaria monitoring frameworks and SDG 3.3 indicators.

### Explanatory variables

Two equity dimensions were used: Wealth Quintiles: Based on the DHS/MICS wealth index, which is constructed using principal component analysis of household assets, housing characteristics, and access to services. The population is categorized into five quintiles: poorest (Q1), poorer (Q2), middle (Q3), richer (Q4), and richest (Q5). Residence: Coded as rural or urban based on the administrative classification used in each national survey.

### Measures of inequalities

To quantify inequality, four standard equity summary measures were computed using WHO’s Health Equity Assessment Toolkit (HEAT Platform):

Absolute Difference (D): The simple arithmetic difference in ITN/IRS coverage between the most advantaged and least advantaged subgroup (e.g., richest vs. poorest, or urban vs. rural).

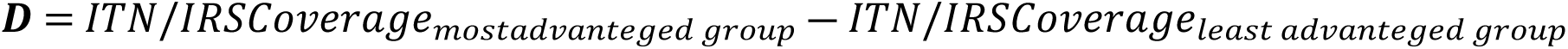

Ratio (R): A relative measure calculated by dividing the coverage rate in the most advantaged group by that of the least advantaged group.

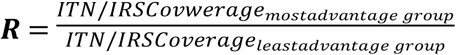

Population Attributable Risk (PAR): Estimates the absolute gain in national coverage that would be achieved if all subgroups had the same coverage as the most advantaged group.

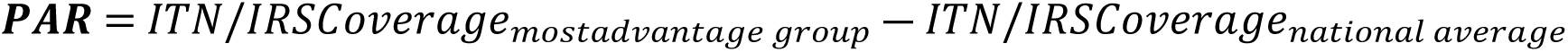

Population Attributable Fraction (PAF): Expresses PAR as a percentage of the national average, indicating the proportion of the national gap that could be eliminated.

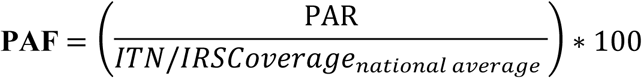

These measures offer both absolute and relative perspectives of inequality and are suitable for monitoring health equity across multiple settings.

### Data Analysis

Data cleaning and initial exploratory analyses, including tabulations and visualization, were conducted using Microsoft Excel. Excel facilitated the organization and preliminary review of household survey data for each country, enabling clear descriptive summaries and preparation for further analysis.

All equity summary measures were computed consistently across countries using the World Health Organization’s Health Equity Assessment Toolkit (HEAT) software. HEAT is a specialised, free application designed to analyze and report health inequalities systematically. It calculates multiple summary measures of inequality such as absolute difference (D), ratio (R), population attributable risk (PAR), and population attributable fraction (PAF), across wealth quintiles and residence (urban versus rural). The software allows for standardized equity analysis using disaggregated data, ensuring comparability and robustness of inequality assessments in the malaria prevention coverage indicators examined.

For geographic visualization, maps illustrating the spatial distribution and patterns of inequality were created using Datawrapper. Country codes compliant with ISO 3166-1 alpha-3 standards were used for accurate and consistent country identification on maps, enhancing clarity in the presentation of regional disparities.

Together, these tools enabled a comprehensive analytical workflow: Excel supported data management and exploratory summaries, WHO HEAT provided rigorous inequality computations, and Datawrapper delivered intuitive visual communication of geographic inequalities in coverage.

## Results

### Coverage of Household Malaria Prevention Measures (ITN and/or IRS)

The analysis revealed substantial variation in national coverage of household-level malaria prevention measures across the 34 African countries. The highest proportions of households with at least one insecticide-treated net (ITN) and/or indoor residual spraying (IRS) in the past 12 months were observed in Mali (93.6%), Benin (92.0%), Burkina Faso (89.8%), Rwanda (89.2%), and Zambia (83.3%). Countries such as Senegal (84.5%), Uganda (84.2%), and Madagascar (80.9%) also reported high levels of coverage. Moderate coverage levels were observed in Ghana (74.1%), Gambia (73.9%), Nigeria (69.0%), and Mozambique (68.7%). On the other end of the spectrum, Ethiopia (3.4%) and Eswatini (4.4%) reported the lowest coverage levels, indicating limited access to key vector control interventions. Other countries with notably low coverage included Angola (31.8%), Namibia (32.7%), Congo (33.1%), and Cameroon (37.6%) as shown in Figure1

### Wealth-Based Inequalities in ITN/IRS Coverage

Table1 & Figure 2: Marked disparities in household ownership of at least one insecticide-treated net (ITN) and/or receipt of indoor residual spraying (IRS) were observed across wealth quintiles. While many countries achieved high coverage across all wealth groups, several exhibited notable pro-poor or pro-rich patterns. In Benin, for instance, coverage was fairly equitable, with uptake ranging from 89.6% among the poorest to 90.4% among the richest, indicating minimal disparity. Burkina Faso also showed consistent high coverage across quintiles (84.4%–93.8%), suggesting successful nationwide implementation. In contrast, some countries showed pro-rich inequality. For example, in Burundi, coverage increased steadily from 30.7% in the poorest quintile to 64.3% in the richest, reflecting pronounced disparities. Similarly, in Mozambique, coverage ranged from 56.0% among the poorest to 80.4% among the richest. However, reverse (pro-poor) inequalities were also present. In Nigeria, the poorest households had higher ITN/IRS coverage (86.2%) compared to the richest (58.0%). A similar trend was observed in Senegal (92.2% poorest vs. 74.3% richest) and Gambia (84.9% poorest vs. 56.3% richest), possibly reflecting targeted interventions in high-burden, low-income areas. Other countries showed more mixed or inconsistent patterns. For example, in Côte d’Ivoire, coverage was relatively high across all quintiles but did not follow a clear gradient (71.8% poorest vs. 61.3% richest). Liberia had a peak in the second quintile (72.1%), with a decline among both the poorest and richest groups.

**Figure 1:**
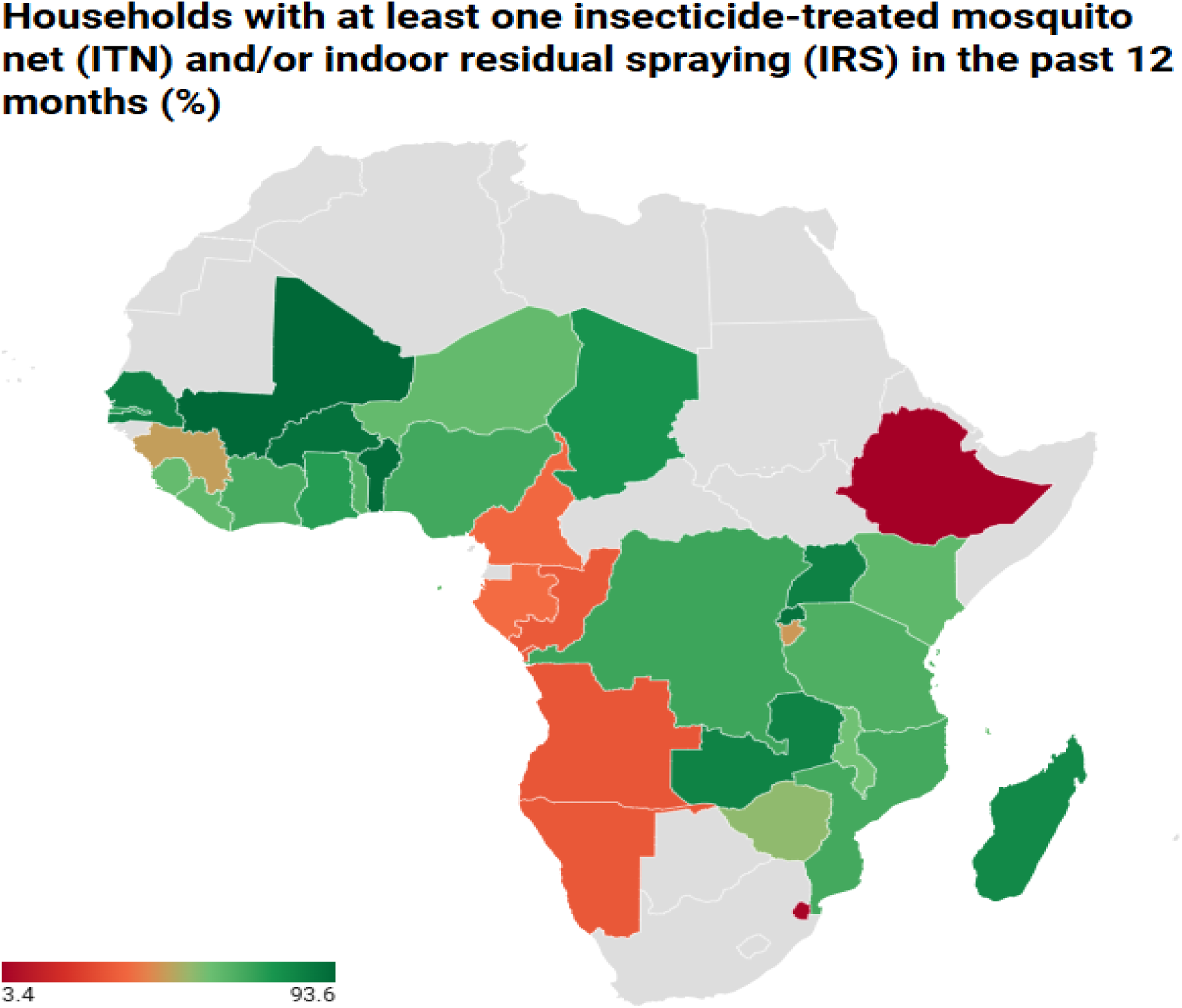
Insecticide-Treated Nets and Indoor Residual Spraying Coverage in West Africa

**Figure 2:**
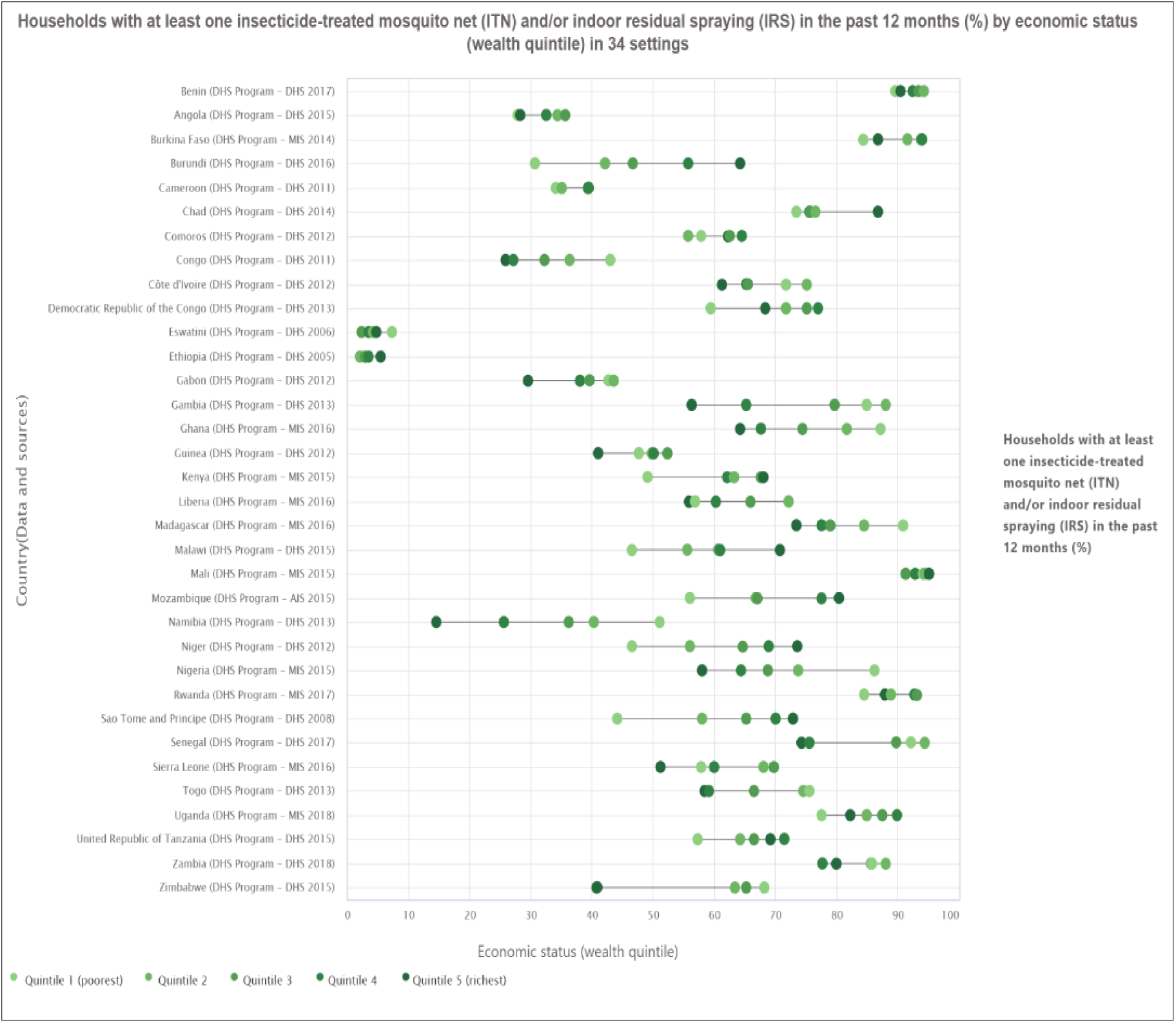
Wealth Inequality in Insecticide-Treated Nets and Indoor Residual Spraying in West Africa

### Residence-Based Inequalities in ITN/IRS Coverage

Table 1 & Figure 3: show substantial differences in household coverage of at least one insecticide-treated mosquito net (ITN) and/or indoor residual spraying (IRS) were observed across rural and urban settings in sub-Saharan Africa. In several countries, rural households reported higher coverage than their urban counterparts. Notably, Benin (93.1% rural vs. 90.5% urban), Burkina Faso (90.8% vs. 87.4%), and Mali (93.4% vs. 94.1%) all demonstrated near-universal coverage in both settings, with a slight rural advantage in most cases. Conversely, some countries recorded markedly higher coverage in urban areas. For example, Burundi reported 64.8% in urban areas compared to 44.5% in rural, and Namibia had 49.7% rural versus 17.0% urban, indicating substantial disparities. Several countries exhibited minimal rural-urban gaps. For instance, Ghana (84.2% rural vs. 65.7% urban) and Kenya (62.8% vs. 62.1%) had relatively balanced coverage, though Ghana still showed a pronounced rural advantage.

**Figure 3:**
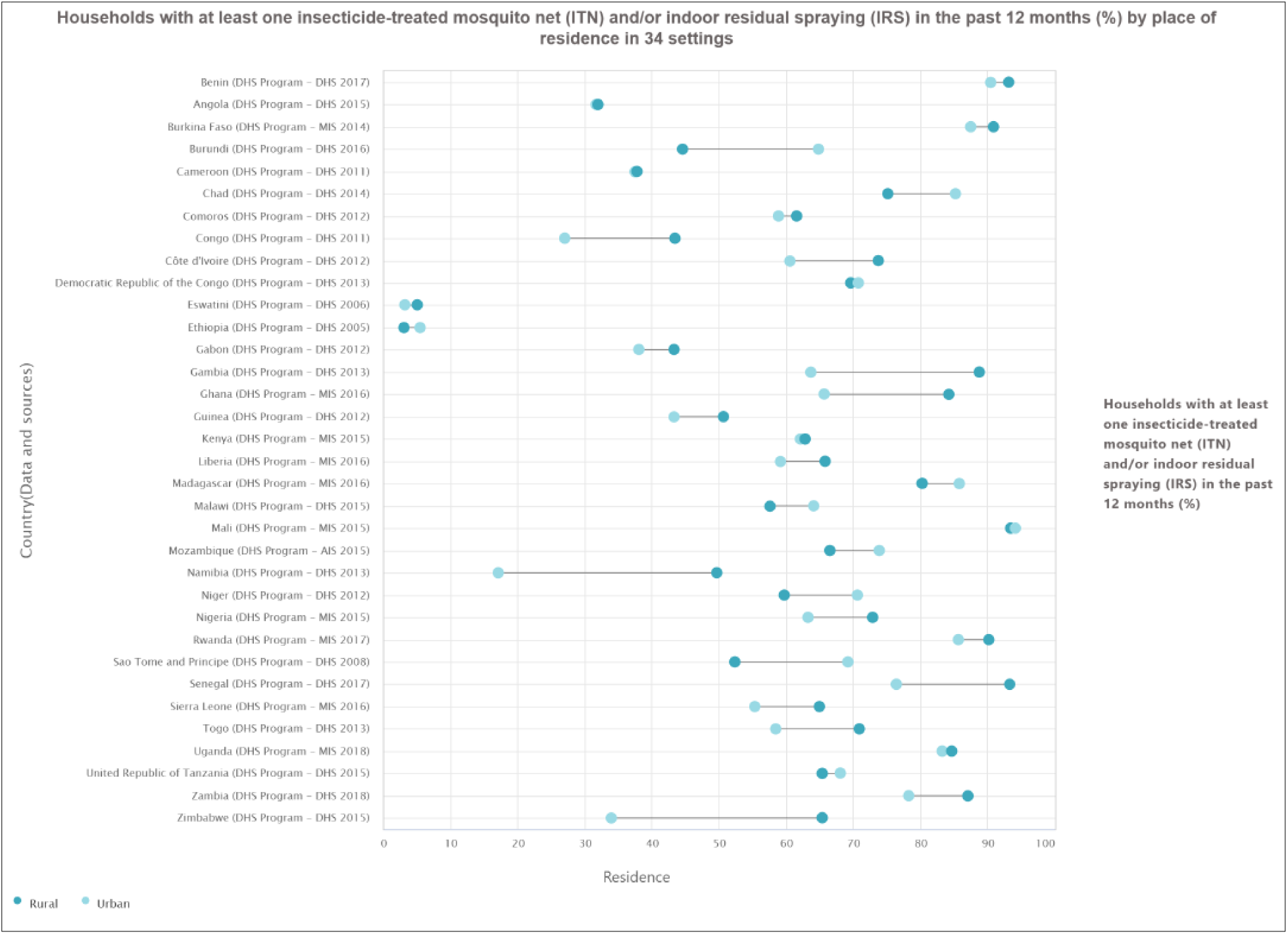
Residential inequality in Insecticide-Treated Nets and Indoor Residual Spraying in West Africa

**Table 1:** Wealth-Based Inequalities in ITN/IRS Coverage.

| Country | Residence |  | Wealth Quintiles |  |  |  |  |
| --- | --- | --- | --- | --- | --- | --- | --- |
|  | Rural | Urban | Quintile 1<br>(poorest) | Quintile 2 | Quintile 3 | Quintile 4 | Quintile 5<br>(richest) |
| Angola | 32.0 | 31.7 | 27.9 | 34.3 | 35.6 | 32.5 | 28.3 |
| Benin | 93.1 | 90.5 | 89.6 | 94.2 | 93.4 | 92.4 | 90.4 |
| Burkina Faso | 90.8 | 87.4 | 84.4 | 91.6 | 93.8 | 94.0 | 86.8 |
| Burundi | 44.5 | 64.8 | 30.7 | 42.2 | 46.7 | 55.8 | 64.3 |
| Côte d'Ivoire | 73.7 | 60.6 | 71.8 | 75.2 | 65.5 | 65.2 | 61.3 |
| Cameroon | 37.7 | 37.5 | 34.1 | 35.1 | 39.3 | 39.5 | 39.4 |
| Chad | 75.1 | 85.2 | 73.4 | 76.6 | 75.7 | 75.6 | 86.7 |
| Comoros | 61.5 | 58.8 | 57.8 | 55.7 | 62.5 | 64.5 | 62.3 |
| Congo | 43.4 | 27.0 | 43.0 | 36.4 | 32.2 | 27.1 | 25.9 |
| DR Congo | 69.6 | 70.7 | 59.4 | 71.7 | 75.2 | 77.0 | 68.4 |
| Eswatini | 5.0 | 3.2 | 7.3 | 4.2 | 2.4 | 3.5 | 4.7 |
| Ethiopia | 3.1 | 5.4 | 2.9 | 2.1 | 2.9 | 3.4 | 5.5 |
| Gabon | 43.3 | 38.1 | 42.7 | 43.5 | 39.6 | 38.0 | 29.6 |
| Gambia | 88.7 | 63.7 | 84.9 | 88.1 | 79.7 | 65.2 | 56.3 |
| Ghana | 84.2 | 65.7 | 87.2 | 81.7 | 74.5 | 67.7 | 64.2 |
| Guinea | 50.6 | 43.3 | 47.6 | 49.8 | 52.4 | 50.1 | 41.0 |
| Kenya | 62.8 | 62.1 | 49.1 | 63.2 | 67.6 | 62.1 | 68.0 |
| Liberia | 65.8 | 59.2 | 56.8 | 72.1 | 66.0 | 60.3 | 55.9 |
| Madagascar | 80.3 | 85.7 | 90.9 | 84.5 | 78.9 | 77.6 | 73.5 |
| Malawi | 57.6 | 64.1 | 46.6 | 55.6 | 60.7 | 61.0 | 70.8 |
| Mali | 93.4 | 94.1 | 94.1 | 94.4 | 91.3 | 92.8 | 95.1 |
| Mozambique | 66.5 | 73.8 | 56.0 | 66.8 | 67.1 | 77.6 | 80.4 |
| Namibia | 49.7 | 17.0 | 51.0 | 40.3 | 36.2 | 25.6 | 14.5 |
| Niger | 59.7 | 70.6 | 46.5 | 56.0 | 64.7 | 68.9 | 73.6 |
| Nigeria | 72.9 | 63.2 | 86.2 | 73.7 | 68.8 | 64.4 | 58.0 |
| Rwanda | 90.1 | 85.6 | 84.5 | 88.9 | 93.2 | 92.7 | 87.9 |
| Sao Tome and Principe | 52.4 | 69.2 | 44.1 | 58.0 | 65.2 | 70.0 | 72.9 |
| Senegal | 93.3 | 76.4 | 92.2 | 94.4 | 89.8 | 75.6 | 74.3 |
| Sierra Leone | 64.9 | 55.3 | 57.9 | 68.0 | 69.8 | 60.0 | 51.2 |
| Togo | 70.9 | 58.4 | 75.5 | 74.6 | 66.5 | 59.1 | 58.4 |
| Uganda | 84.6 | 83.2 | 77.5 | 84.9 | 87.5 | 89.9 | 82.2 |
| Tanzania | 65.3 | 68.1 | 57.3 | 64.2 | 66.5 | 71.5 | 69.2 |
| Zambia | 87.1 | 78.2 | 85.7 | 88.1 | 85.6 | 77.7 | 80.0 |
| Zimbabwe | 65.4 | 33.9 | 68.2 | 63.4 | 65.2 | 40.7 | 40.9 |

### Wealth-Related Inequality Measures in Household Access to ITN/IRS across 34 African Countries

Table 2 revealed wealth-related disparities in household access to at least one insecticide-treated mosquito net (ITN) and/or indoor residual spraying (IRS) within the past 12 months varied markedly across the 33 African countries examined. Substantial absolute differences (D) in coverage between the poorest and richest households were observed. The most pronounced pro-rich gaps were recorded in Burundi (33.6 percentage points), São Tomé and Príncipe (28.8), Niger (27.1), Mozambique (24.4), and Malawi (24.2), indicating significantly higher access among the wealthiest groups. In contrast, countries such as Namibia (–36.5), Gambia (–28.6), Nigeria (–28.2), Zimbabwe (–27.3), and Ghana (–23.0) displayed strong pro-poor gradients, where poorer households reported higher access than their richer counterparts.

**Table 2:** Wealth-Related Inequality Measures in Household Access to ITN/IRS across Selected African Countries.

| Country | Year | D | R | PAF | PAR |
| --- | --- | --- | --- | --- | --- |
| Angola | 2015 | 0.4 | 1.0 | 0.0 | 0.0 |
| Benin | 2015 | 0.8 | 1.0 | 0.0 | 0.0 |
| Burkina Faso | 2014 | 2.4 | 1.0 | 0.0 | 0.0 |
| Burundi | 2016 | 33.6 | 2.1 | 37.5 | 17.5 |
| Côte d'Ivoire | 2016 | -10.5 | 0.9 | 0.0 | 0.0 |
| Cameroon | 2011 | 5.3 | 1.2 | 4.8 | 1.8 |
| Chad | 2014 | 13.3 | 1.2 | 12.2 | 9.4 |
| Comoros | 2012 | 4.5 | 1.1 | 2.9 | 1.7 |
| Congo | 2011 | -17.1 | 0.6 | 0.0 | 0.0 |
| DR Congo | 2013 | 9.0 | 1.2 | 0.0 | 0.0 |
| Eswatini | 2006 | -2.6 | 0.6 | 7.2 | 0.3 |
| Ethiopia | 2005 | 2.6 | 1.9 | 62.7 | 2.1 |
| Gabon | 2012 | -13.1 | 0.7 | 0.0 | 0.0 |
| Gambia | 2013 | -28.6 | 0.7 | 0.0 | 0.0 |
| Ghana | 2016 | -23.0 | 0.7 | 0.0 | 0.0 |
| Guinea | 2012 | -6.6 | 0.9 | 0.0 | 0.0 |
| Kenya | 2015 | 18.9 | 1.4 | 8.8 | 5.5 |
| Liberia | 2016 | -0.9 | 1.0 | 0.0 | 0.0 |
| Madagascar | 2016 | -17.4 | 0.8 | 0.0 | 0.0 |
| Malawi | 2015 | 24.2 | 1.5 | 20.8 | 12.2 |
| Mali | 2015 | 1.0 | 1.0 | 1.7 | 1.5 |
| Mozambique | 2015 | 24.4 | 1.4 | 17.0 | 11.7 |
| Namibia | 2013 | -36.5 | 0.3 | 0.0 | 0.0 |
| Niger | 2012 | 27.1 | 1.6 | 19.7 | 12.1 |
| Nigeria | 2015 | -28.2 | 0.7 | 0.0 | 0.0 |
| Rwanda | 2017 | 3.4 | 1.0 | 0.0 | 0.0 |
| Sao Tome and Principe | 2008 | 28.8 | 1.7 | 20.0 | 12.1 |
| Senegal | 2017 | -17.9 | 0.8 | 0.0 | 0.0 |
| Sierra Leone | 2016 | -6.7 | 0.9 | 0.0 | 0.0 |
| Togo | 2013 | -17.1 | 0.8 | 0.0 | 0.0 |
| Uganda | 2018 | 4.7 | 1.1 | 0.0 | 0.0 |
| Tanzania | 2015 | 11.9 | 1.2 | 4.5 | 3.0 |
| Zambia | 2018 | -5.7 | 0.9 | 0.0 | 0.0 |
| Zimbabwe | 2015 | -27.3 | 0.6 | 0.0 | 0.0 |

The ratio (R) similarly highlighted the magnitude of disparities. In Burundi, access among the richest was more than twice that of the poorest (R = 2.1), while São Tomé and Príncipe (R = 1.7), Niger (R = 1.6), Malawi (R = 1.5), and Mozambique (R = 1.4) showed substantial relative pro-rich inequality. Meanwhile, strong pro-poor trends were reflected in Namibia (R = 0.3), Zimbabwe (0.6), Gambia (0.7), Nigeria (0.7), and Ghana (0.7), where the relative likelihood of ITN/IRS access favored the poorest households.

The population attributable risk (PAR), which estimates the potential increase in national coverage if all socioeconomic groups had the same access level as the richest, was highest in Burundi (17.5 percentage points), followed by Malawi (12.2), Niger and São Tomé and Príncipe (both 12.1), and Mozambique (11.7). These values suggest that addressing inequality in these countries could yield considerable national gains in malaria prevention coverage.

Similarly, the population attributable fraction (PAF) representing the proportion of the national coverage shortfall due to inequality was highest in Ethiopia (62.7%), Burundi (37.5%), Malawi (20.8%), Niger (19.7%), and São Tomé and Príncipe (20.0%). In contrast, most countries with pro-poor gradients (e.g., Nigeria, Ghana, and Zimbabwe) recorded negligible or zero PAF values, indicating minimal or no inequality-attributable loss in national coverage.

### *Residence-*Related Inequality Measures in Household Access to ITN/IRS across 34 African Countries

Table 3 shows marked disparities in household access to ITN and/or IRS across rural and urban areas, with the direction and magnitude of inequality varying significantly by country. In terms of absolute difference (D), countries such as Burundi (20.3), São Tomé and Príncipe (16.8), Niger (10.9), and Mozambique (7.3) showed a pronounced urban advantage in ITN/IRS coverage. While Zimbabwe (–31.5 percentage points), Namibia (–32.7), Ghana (–18.5), and Senegal (–16.9) reported significantly higher coverage among rural households than their urban counterparts.

**Table 3:** Residence-Related Inequality Measures in Household Access to ITN/IRS across 34 African Countries.

| Country | Year | D | R | PAF | PAR |
| --- | --- | --- | --- | --- | --- |
| Angola | 2015 | -0.3 | 1.0 | 0.0 | 0.0 |
| Benin | 2015 | -2.6 | 1.0 | 0.0 | 0.0 |
| Burkina Faso | 2014 | -3.4 | 1.0 | 0.0 | 0.0 |
| Burundi | 2016 | 20.3 | 1.5 | 38.7 | 18.1 |
| Côte d'Ivoire | 2016 | -13.1 | 0.8 | 0.0 | 0.0 |
| Cameroon | 2011 | -0.2 | 1.0 | 0.0 | 0.0 |
| Chad | 2014 | 10.1 | 1.1 | 10.3 | 8.0 |
| Comoros | 2012 | -2.7 | 1.0 | 0.0 | 0.0 |
| Congo | 2011 | -16.4 | 0.6 | 0.0 | 0.0 |
| DR Congo | 2013 | 1.1 | 1.0 | 1.1 | 0.8 |
| Eswatini | 2006 | -1.8 | 0.6 | 0.0 | 0.0 |
| Ethiopia | 2005 | 2.3 | 1.7 | 57.4 | 2.0 |
| Gabon | 2012 | -5.2 | 0.9 | 0.0 | 0.0 |
| Gambia | 2013 | -25.0 | 0.7 | 0.0 | 0.0 |
| Ghana | 2016 | -18.5 | 0.8 | 0.0 | 0.0 |
| Guinea | 2012 | -7.3 | 0.9 | 0.0 | 0.0 |
| Kenya | 2015 | -0.7 | 1.0 | 0.0 | 0.0 |
| Liberia | 2016 | -6.6 | 0.9 | 0.0 | 0.0 |
| Madagascar | 2016 | 5.4 | 1.1 | 5.9 | 4.8 |
| Malawi | 2015 | 6.5 | 1.1 | 9.4 | 5.5 |
| Mali | 2015 | 0.7 | 1.0 | 0.6 | 0.5 |
| Mozambique | 2015 | 7.3 | 1.1 | 7.4 | 5.1 |
| Namibia | 2013 | -32.7 | 0.3 | 0.0 | 0.0 |
| Niger | 2012 | 10.9 | 1.2 | 14.8 | 9.1 |
| Nigeria | 2015 | -9.7 | 0.9 | 0.0 | 0.0 |
| Rwanda | 2017 | -4.5 | 1.0 | 0.0 | 0.0 |
| Sao Tome and Principe | 2008 | 16.8 | 1.3 | 13.9 | 8.5 |
| Senegal | 2017 | -16.9 | 0.8 | 0.0 | 0.0 |
| Sierra Leone | 2016 | -9.6 | 0.9 | 0.0 | 0.0 |
| Togo | 2013 | -12.5 | 0.8 | 0.0 | 0.0 |
| Uganda | 2018 | -1.4 | 1.0 | 0.0 | 0.0 |
| Tanzania | 2015 | 2.8 | 1.0 | 2.8 | 1.9 |
| Zambia | 2018 | -8.9 | 0.9 | 0.0 | 0.0 |
| Zimbabwe | 2015 | -31.5 | 0.5 | 0.0 | 0.0 |

Relative inequality assessed by the ratio (R) revealed substantial pro-urban disparities in a few countries. Burundi (R = 1.5), Ethiopia (1.7), São Tomé and Príncipe (1.3), and Niger (1.2) all exhibited R values above 1.0, indicating higher access in urban areas. In contrast, countries such as Zimbabwe (R = 0.5), Namibia (0.3), and Congo (0.6) recorded strong pro-rural gradients, with rural households more likely to have access to ITN/IRS than urban households.

In terms of potential national gains, Burundi had the highest population attributable risk (PAR) at 18.1 percentage points, followed by Niger (9.1), São Tomé and Príncipe (8.5), and Chad (8.0). This suggests that harmonizing rural and urban coverage at the level of the most advantaged subgroup could yield notable improvements in national ITN/IRS coverage. Finally, the population attributable fraction (PAF) representing the percentage of total inequality in coverage that could be addressed was highest in Ethiopia (57.4%), Burundi (38.7%), Niger (14.8%), and São Tomé and Príncipe (13.9%).

## Discussion

### Overall ITN/IRS Coverage Levels

There were stark cross-country differences in the study findings concerning coverage of household malaria prevention measures, ranging from near-universal coverage in some to almost non-existent in others. In the best-performing countries, such as Mali, Benin, Burkina Faso, Rwanda, Zambia, and Senegal, more than 80% of households had either one ITN or a recent instance of IRS, with Mali leading at 93.6%. These countries have defied frontal scaling-up of vector control to actualise control in most households. Especially those of the Sahel countries (for instance, Mali, Burkina Faso) have benefited from periodic mass ITN campaigns bolstered by international donors and strong national will for malaria control (14,15).

These achievements align with other reports presenting these countries as positive outliers in ITN coverage. As an example, in recent World Malaria Reports, Burkina Faso and Rwanda were identified as having rapidly scaled up net distributions in the 2010s, which led to the increased net ownership at the household level (14). High net coverage at the household level is a strong positive sign that universal coverage strategies, such as free LLIN distribution through campaigns and continuous channels, have been applied well. A number of these high-coverage countries also witnessed relatively equitable coverage between subgroups (discussed further below), providing evidence that large-scale deployments can be balanced with equity (23).

Some countries, on the contrary, revealed a very low household access to ITN/IRS, with less than 40% coverage. Ethiopia (3.4%) and Eswatini (4.4%) stood at the very bottom. In these extreme cases, fewer than 1 in 20 homes are protected from malaria, and this merits careful interpretation. Ethiopian survey data (2015 in this analysis) must be interpreted in the light of the fact that historically, certain areas relied heavily on IRS. In contrast, ITN coverage at the national level was extremely low (24). Nationwide ITN campaigns did not start until very late, around the late 2000s, and there are many areas within Ethiopia above the highlands, where the risk of malaria is relatively low. Hence, more contemporary Malaria Indicator Surveys have since reported higher coverage, at least following targeted distributions to the endemic regions, yet sizeable challenges continue in maintaining coverage given the population size and ecological diversity (24)

Eswatini’s very low coverage is a probable testimony to the context of near elimination. By 2017, the country was recording barely a few hundred malaria cases annually, having transitioned from mass campaigns for IRS to targeted IRS and surveillance (15). Other countries with considerably low coverage include Angola, Namibia, Congo, and Cameroon (each with about 30–37% coverage). Such low levels are particularly worrisome in high-burden contexts like Angola, which, together with Nigeria, accounts for some of the highest mortality rates attributed to malaria in the world (15), such a discrepancy in need and coverage points to programmatic gaps. Conflict, weak health systems, and logistical challenges often interrupt ITN/IRS delivery in these scenarios (23). For instance, Nigeria (with 69% coverage in our results) has distributed more than 100 million ITNs in the last decade. However, areas remain uncovered due to issues such as retention of nets by the communes and terms of accessibility (23).

This variation in national coverage thus points to the role of contextual factors such as political commitment, partner support, infrastructure, and stability in defining the outcome. Those with a heavy malaria burden but who are now off track (e.g., Angola, Cameroon) may require heightened assistance and new delivery paradigms. Meanwhile, the high performers serve as the exemplars for success but need to protect these gains through regular net replacement and annual IRS campaigns (13).

### Wealth-Related Inequalities

One of the study’s findings is that in some countries, household ITN/IRS access exhibits significant disparities along wealth lines, whereas in others, there are none or even a reversal. At one extreme, a handful of countries show larger pro-rich inequalities. Burundi records the most considerable wealth-based disparity: coverage in the richest wealth quintile exceeds the poorest by 33.6 percentage points, with the coverage ratio (R) standing at 2.1. São Tomé and Príncipe (28.8 pp, R =1.7), Niger (27.1 pp, R=1.6), Mozambique (24.4 pp, R=1.4), and Malawi (24.2 pp, R=1.5) followed very similar patterns. So, it is indicative that the poorest households remain underserved by vector control interventions (25).

Such pro-rich inequities might stem from distribution methods for nets that fail to reach the most deprived. In Niger and Mozambique, however, depending mostly on periodic campaigns without continuous distribution may leave at least some poor and remote communities vulnerable to malaria between campaign rounds (26). In Malawi, the emergence of private IRS among the richer may aggravate the existing inequality in IRS coverage because the poor rely solely on ITNs supplied publicly (25). These patterns corroborate earlier studies that socioeconomic status is the primary determinant of ITN ownership when distribution is incomplete (27). Furthermore, socioeconomic inequality has consistently been found in the uptake of major malaria interventions such as intermittent preventive treatment in pregnancy (IPTp) across many African countries, pointing to a systemic challenge in reaching the most vulnerable (28,29).

In contrast, the study has shown pro-poor coverage in some countries: Namibia (–36.5 pp), Gambia (–28.6 pp), Nigeria (–28.2 pp), Zimbabwe (–27.3 pp), and Ghana (–23.0 pp), with negative coverage gaps showing greater coverage among the poorest. In Nigeria, targeted door-to-door ITN campaigns supported by the Global Fund and PMI prioritized rural northern states with intense malaria transmission and high poverty (23,24). A similar rationale accounts for the results in Zimbabwe and Ghana, where net distribution has been focused in rural malaria zones with high poverty levels (23). These results for a pro-poor bias are mainly given by mention, as the programs were intentionally designed to direct resources to areas of most significant burden and fewest resources, thus upholding the principles of equity from theory to practical public health impact.

Encouragingly, in nearly half of the countries studied, wealth-related inequalities were minimal (D = 0, R =1). This suggests that well-executed universal coverage campaigns can generate equity, regardless of wealth status. Previous research, similarly, noted that by 2015, roughly half of African countries had no detectable wealth gap in ITN access (25). This stresses that with willful efforts, critical malaria interventions can be accessed equitably.

From a policy perspective, these inequality patterns carry very tangible implications. Using our population-attributable risk (PAR) and fraction (PAF) estimates clearly illustrates how much national coverage might be increased if disparities were removed. For instance, Burundi’s PAR of 17.5 percentage points implies a significant increase in household protection if parity between the rich and poor can be brought about. Ethiopia’s PAF of 62.7% indicates that nearly two-thirds of its gap is driven by inequalities. On the other hand, pro-poor Nigeria and Ghana, with practically zero PAF, show inequality is no longer a barrier in these countries (30). Such figures give a quantitative meaning to the idea of promoting interventions that lessen inequities.

### Residence-Based Inequality

The study has put emphasis, besides wealth, on the urban-rural divide as it relates to the opportunity of owning ITNs/IRS. In some countries, urban households are more likely to possess ITNs/IRS (Burundi, Ethiopia, São Tomé and Príncipe, Niger, Chad), possibly due to early distribution through health facilities in urban areas or the challenges faced in distribution in rural areas (WHO, 2024b). Some of the data from Ethiopia in 2005 point to this early distribution phase(24). Such pro-urban biases become problematic as malaria transmission is generally high in rural areas where access to health and prevention tools are somewhat difficult (31). Malaria knowledge and perceived intervention efficacy also matter in uptake and are known to be different between urban and rural areas (32).

Conversely, a few countries demonstrated pro-rural scenarios: Namibia, Zimbabwe, Gambia, Ghana, and Nigeria. These findings can be explained by malaria program strategies that favor rural high-transmission areas. In Ghana, for example, rural net ownership was higher than urban net ownership because of national targeting (23). These findings suggest that, at least in some countries, malaria control tools have reached most rural poor, which is a positive indicator in terms of equity (25). What happened in these countries shows very well that reaching underserved rural populations beyond geographical barriers is possible with targeted and flexible delivery mechanisms, including community-based ones (González et al., 2023; Koita et al., 2024).

Where no urban-rural differences were observed at least to a significant extent (e.g., Kenya, Mali, Senegal), the deliveries appeared to be pretty balanced between urban and rural settings. In countries where some residual pro-urban gaps are observed, the need for strengthened rural engagement, including through CHWs and mobile distribution, therefore remains (15,35). These results also highlight the need for understanding the specific roots of inequality in each country to design context-specific and truly effective interventions.

### Malaria Control and Equity: Implications

These findings suggest that equity is important for both the moral and pragmatic aspects of malaria control. Any existing pro-rich or pro-urban bias allows the most vulnerable populations to remain susceptible to malaria transmission, thereby hampering the practical achievement of the 2025 goals of malaria control (23,31). Essentially, equitable distributions of ITNs and IRS are the fundamental requirements for universal health coverage and sustainable disease control. The findings of this study offer updates for national malaria control programs, African Union initiatives, and other international partners to revise strategies that will maximize interventions for greater impact (5).

Societal policies should consider the following: targeting underserved populations using, for example, the WHO Health Equity Assessment Toolkit (36); tracking inequality through the regular analysis of DHS/MIS data; and embedding equity into program planning, such as engaging in sub-national equity goal-setting. Kenya and Mali provide examples of countries where equity is a reality (25). Moreover, community-based approaches that have improved IPTp coverage by overcoming some access barriers offer valuable lessons for ITN and IRS distribution (33,34,37). By placing equity at the heart of decision-making, states can thus speed malaria elimination and serve the cause of general health equity on the continent.

### Limitation

This study employed a binary measure of household malaria prevention coverage, defined as having at least one insecticide-treated net (ITN) or recent indoor residual spraying (IRS). While this metric does not capture adequate coverage for all household members, nor does it assess actual net usage or IRS quality, it remains a widely used and accepted indicator for monitoring malaria control progress across countries. Consequently, our findings provide a valuable overview of cross-country patterns and inequalities in access to vector control tools.

We acknowledge that relying on this binary indicator may overestimate functional protection and limits detailed understanding of the drivers behind observed disparities. Moreover, the survey data utilized spans from 2005 to 2018, which may not fully reflect more recent programmatic advances or changes in malaria transmission dynamics. Nonetheless, these datasets remain some of the most comprehensive and comparable sources for assessing population-level equity in malaria prevention across sub-Saharan Africa.

Despite these limitations, the study significantly contributes to the literature by systematically documenting wealth- and residence-based inequalities in access to malaria prevention interventions at a continental scale. These insights offer policymakers critical information for prioritizing equity-driven malaria control efforts. We recommend that future research incorporate more granular and recent data on adequate net coverage, usage, and intervention quality to deepen understanding and strengthen targeted strategies.

## Conclusion

This study highlights continued and significant inequalities in household access to insecticide-treated nets (ITNs) and indoor residual spraying (IRS) across 34 sub-Saharan African countries. Even though some countries are achieving high overall coverage, marked disparities remain both socioeconomically and geographically. Wealth-related inequalities varied widely, with some countries displaying pronounced pro-rich access while others evidenced pro-poor patterns. This required differentiated targeting and implementation strategies within national malaria control programs. Also, rural-urban disparities were persisted; in many settings, rural households had higher coverage, although urban advantages were evident elsewhere.

Importantly, inequality metrics including absolute difference, ratio, population attributable risk (PAR), and population attributable fraction (PAF) underscored the potential national gains from addressing these disparities. Countries such as Burundi, Malawi, and São Tomé and Príncipe where inequalities substantially depress overall malaria prevention coverage. The findings suggest that equity-focused strategies that prioritize underserved populations and settings could significantly enhance the effectiveness and reach of ITN and IRS interventions.

In pursuit of malaria elimination and attainment of Sustainable Development Goal 3.3, targeted policies and resource allocation must focus on closing these gaps to ensure no population groups are left behind. Strengthening data monitoring systems for equity, improving the distribution channels, and tailoring interventions to local epidemiological and socioeconomic contexts are critical for optimising vector control impact. Ultimately, bridging these inequalities will be essential to achieve universal coverage and reduce the malaria burden equitably across sub-Saharan Africa.

## Data Availability

The datasets generated and/or analyzed during this study are available in the WHO's HEAT version 5.0: https://www.who.int/data/inequality-monitor/data

## Acknowledgements

We express our gratitude to the WHO for providing access to HEAT software free of charge to the public.

## Abbreviations

ITN: Insecticide-Treated
Net IRS: Indoor Residual Spraying
DHS: Demographic and Health Surveys
MIS: Malaria Indicator Surveys
MICS: Multiple Indicator Cluster Surveys
WHO: World Health Organization
HEM: Health Equity Monitor
GHO: Global Health Observatory
SDG: Sustainable Development Goal
PAR: Population Attributable Risk
PAF: Population Attributable Fraction
R: Ratio (relative inequality measure)
D: Absolute Difference (absolute inequality measure)

## Ethics approval and consent to participate

Ethics approval was not needed, as the data were available to the public.

## Consent for publication

Not applicable.

## Authors’ contributions

**AI** and **SZ** conceived the study. **AI, GC**, **TJ, and WA** wrote the methods section and performed the data analysis. **AI, GC, TJ,** were responsible for the initial draft of the manuscript. All the authors reviewed and approved the final version of the manuscript.

## Funding

This study did not receive any funding.

## Competing interests

The authors declare no competing interests.

## Appendix

**Figure 1:**
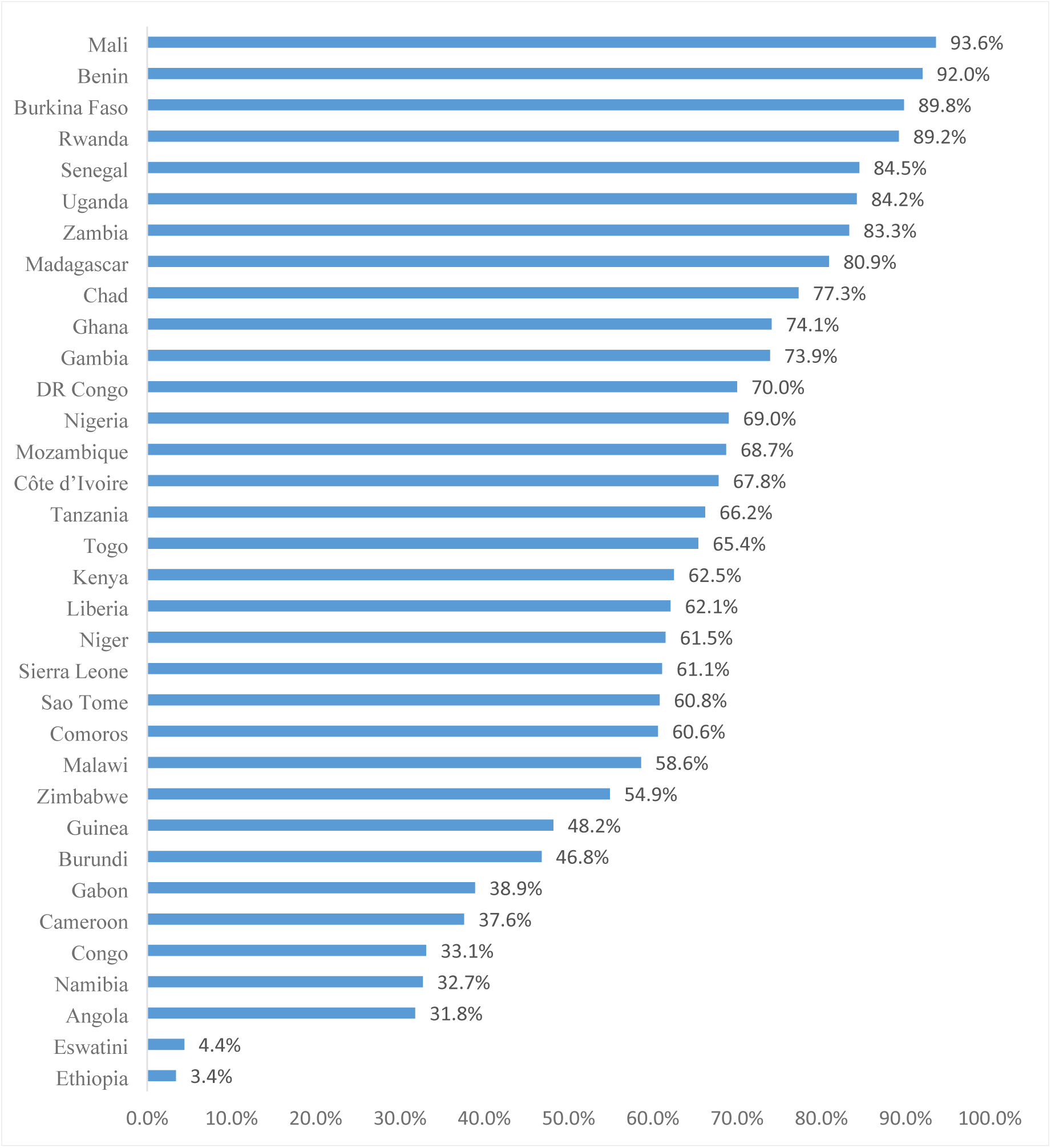
Percentage of Households with at least one (ITN) and/or (IRS) in the past 12 months (%)

## Notes

### Competing Interest Statement

The authors have declared no competing interest.

### Funding Statement

This study did not recieve any funding

### Author Declarations

Ethics approval was not needed, as the data were available to the public.

## Reference

1. WHO. World Health Organization. 2023 [cited 2025 Aug 3]. World malaria report 2023 - World Health Organization (WHO). Available from: https://www.who.int/teams/global-malaria-programme/reports/world-malaria-report-2023

2. WHO. World Health Organization. 2015 [cited 2025 Aug 3]. World malaria report 2015. Available from: http://www.who.int/malaria/visual-refresh/en/

3. WHO. World Health Organization. 2020. World malaria report 2020: 20 years of global progress and challenges.

4. Li J, Docile HJ, Fisher D, Pronyuk K, Zhao L. Current Status of Malaria Control and Elimination in Africa: Epidemiology, Diagnosis, Treatment, Progress and Challenges. J Epidemiol Glob Health [Internet]. 2024 Sep 1 [cited 2025 Aug 3];14(3):561–79. Available from: https://pubmed.ncbi.nlm.nih.gov/38656731/

5. ALMA. African Leaders Malaria Alliance (ALMA. 2024 [cited 2025 Aug 3]. 2024 Africa Malaria Progress Report - African Leaders Malaria Alliance. Available from: https://alma2030.org/heads-of-state-and-government/african-union-malaria-progress-reports/2024-africa-malaria-progress-report/

6. WHO. World Health Organization. 2021 [cited 2025 Aug 3]. Global Malaria Programme. Consolidated guidelines for malaria. Available from: https://www.who.int/teams/global-malaria-programme/guidelines-for-malaria

7. WHO. World Health Organization. 2024 [cited 2025 Aug 3]. World malaria report 2024. Available from: https://www.who.int/teams/global-malaria-programme/reports/world-malaria-report-2024

8. WHO. World Health Organization. 2019. The E-2020 initiative of 21 malaria-eliminating countries: 2019 progress report.

9. Pryce J, Medley N, Choi L. Indoor residual spraying for preventing malaria in communities using insecticide-treated nets. Cochrane Database of Systematic Reviews [Internet]. 2022 Jan 17 [cited 2025 Aug 3];2022(1). Available from: https://pubmed.ncbi.nlm.nih.gov/35038163/

10. Ssempiira J, Nambuusi B, Kissa J, Agaba B, Makumbi F, Kasasa S, et al. Geostatistical modelling of malaria indicator survey data to assess the effects of interventions on the geographical distribution of malaria prevalence in children less than 5 years in Uganda. PLoS One [Internet]. 2017 Apr 1 [cited 2025 Aug 3];12(4):e0174948. Available from: https://journals.plos.org/plosone/article?id=10.1371/journal.pone.0174948

11. Scott J, Kanyangarara M, Nhama A, Macete E, Moss WJ, Saute F. Factors associated with use of insecticide-treated net for malaria prevention in Manica District, Mozambique: a community-based cross-sectional survey. Malar J [Internet]. 2021 Dec 1 [cited 2025 Aug 3];20(1):1–9. Available from: https://malariajournal.biomedcentral.com/articles/10.1186/s12936-021-03738-7

12. Hemingway J, Shretta R, Wells TNC, Bell D, Djimdé AA, Achee N, et al. Tools and Strategies for Malaria Control and Elimination: What Do We Need to Achieve a Grand Convergence in Malaria? PLoS Biol [Internet]. 2016 Mar 2 [cited 2025 Aug 3];14(3):e1002380. Available from: https://journals.plos.org/plosbiology/article?id=10.1371/journal.pbio.1002380

13. WHO. World Health Organization. 2015 [cited 2025 Aug 3]. WHO commends the Roll Back Malaria Partnership’s contribution to global progress as governing board disbands secretariat. Available from: https://www.who.int/news/item/25-08-2015-who-commends-the-roll-back-malaria-partnership-s-contribution-to-global-progress-as-governing-board-disbands-secretariat

14. WHO. World Health Organization. 2024 [cited 2025 Aug 3]. WHO guidelines for malaria, 30 November 2024. Available from: https://www.who.int/teams/global-malaria-programme

15. WHO. World Health Organization. 2024 [cited 2025 Aug 3]. World malaria report 2024. Available from: https://www.who.int/teams/global-malaria-programme/reports/world-malaria-report-2024

16. Ezezika O, El-Bakri Y, Nadarajah A, Barrett K. Implementation of insecticide-treated malaria bed nets in Tanzania: a systematic review. J Glob Health Rep [Internet]. 2022 [cited 2025 Aug 3];6:e2022036. Available from: 10.29392/001c.37363

17. Carshon-Marsh R, Di Ruggiero E. Improving the utilization of insecticide-treated nets for malaria prevention among pregnant women, lactating mothers and children in Sierra Leone: a commentary. Malaria Journal [Internet]. 2025 Dec 1 [cited 2025 Aug 3];24(1):1–6. Available from: https://malariajournal.biomedcentral.com/articles/10.1186/s12936-025-05429-z

18. Terefe B, Habtie A, Chekole B. Insecticide-treated net utilization and associated factors among pregnant women in East Africa: evidence from the recent national demographic and health surveys, 2011–2022. Malar J [Internet]. 2023 Dec 1 [cited 2025 Aug 3];22(1):1–9. Available from: https://malariajournal.biomedcentral.com/articles/10.1186/s12936-023-04779-w

19. Zhou Y, Zhang WX, Tembo E, Xie MZ, Zhang SS, Wang XR, et al. Effectiveness of indoor residual spraying on malaria control: a systematic review and meta-analysis. Infect Dis Poverty. 2022 Dec 1;11(1).

20. WHO. World Health Organization. 2025 [cited 2025 Aug 3]. World health statistics 2025: monitoring health for the SDGs, sustainable development goals. Available from: https://www.who.int/publications/i/item/9789240110496

21. Apeagyei AE, Patel NK, Cogswell I, O’Rourke K, Tsakalos G, Dieleman J. Examining geographical inequalities for malaria outcomes and spending on malaria in 40 malaria-endemic countries, 2010–2020. Malar J [Internet]. 2024 Dec 1 [cited 2025 Aug 3];23(1):1–11. Available from: https://malariajournal.biomedcentral.com/articles/10.1186/s12936-024-05028-4

22. Global Fund. The Global Fund to Fight AIDS, Tuberculosis and Malaria: Strategic Review 2023 (SR2023). 2024.

23. The Global Fund. The Global Fund. 2024 [cited 2025 Aug 4]. Results Report 2024 - The Global Fund to Fight AIDS, Tuberculosis and Malaria. Available from: https://www.theglobalfund.org/en/results/

24. The DHS Program. Demographic and Health Survey Program. 2021 [cited 2025 Aug 4]. The DHS Program - Malaria Publications. Available from: https://dhsprogram.com/topics/malaria-Corner/publications.cfm

25. Galactionova K, Smith TA, de Savigny D, Penny MA. State of inequality in malaria intervention coverage in sub-Saharan African countries. BMC Med [Internet]. 2017 Oct 18 [cited 2025 Aug 3];15(1):1–11. Available from: https://bmcmedicine.biomedcentral.com/articles/10.1186/s12916-017-0948-8

26. Zemene E, Koepfli C, Tiruneh A, Yeshiwondim AK, Seyoum D, Lee MC, et al. Detection of foci of residual malaria transmission through reactive case detection in Ethiopia. Malar J [Internet]. 2018 Oct 26 [cited 2025 Aug 3];17(1):1–10. Available from: https://malariajournal.biomedcentral.com/articles/10.1186/s12936-018-2537-5

27. Pulford J, Saweri OPM, Jeffery C, Siba PM, Mueller I, Hetzel MW. Does test-based prescription of evidence-based treatment for malaria improve treatment seeking and satisfaction? Findings of repeated cross-sectional surveys in Papua New Guinea. BMJ Glob Health [Internet]. 2018 [cited 2025 Aug 3];3:915. Available from: https://gh.bmj.com

28. Muhammad FM, Majdzadeh R, Nedjat S, Sajadi HS, Parsaeian M. Socioeconomic inequality in intermittent preventive treatment using Sulphadoxine pyrimethamine among pregnant women in Nigeria. BMC Public Health [Internet]. 2020 Dec 1 [cited 2025 Aug 3];20(1):1–9. Available from: https://bmcpublichealth.biomedcentral.com/articles/10.1186/s12889-020-09967-w

29. Nishan MDNH, Akter K. Coverage and determinants of Intermittent Preventive Treatment in pregnancy (IPTp) in Cameroon, Guinea, Mali, and Nigeria. PLoS One [Internet]. 2024 Nov 1 [cited 2025 Aug 3];19(11):e0313087. Available from: https://journals.plos.org/plosone/article?id=10.1371/journal.pone.0313087

30. WHO. World Health Organization. 2021 [cited 2025 Aug 3]. Global technical strategy for malaria 2016–2030, 2021 update. Available from: https://www.who.int/publications/i/item/9789240031357

31. WHO. World Health Organization. 2021 [cited 2025 Aug 3]. p. 40 Global Tecnichal Strategy for Malaria. Available from: https://www.who.int/publications/i/item/9789240031357

32. Nganda RY, Drakeley C, Reyburn H, Marchant T. Knowledge of malaria influences the use of insecticide treated nets but not intermittent presumptive treatment by pregnant women in Tanzania. Malar J [Internet]. 2004 Nov 12 [cited 2025 Aug 3];3(1):1–7. Available from: https://malariajournal.biomedcentral.com/articles/10.1186/1475-2875-3-42

33. Koita K, Kayentao K, Worrall E, Van Eijk AM, Hill J. Community-based strategies to increase coverage of intermittent preventive treatment of malaria in pregnancy with sulfadoxine–pyrimethamine in sub-Saharan Africa: a systematic review, meta-analysis, meta-ethnography, and economic assessment. Lancet Glob Health [Internet]. 2024 Sep 1 [cited 2025 Aug 3];12(9):e1456–69. Available from: https://pubmed.ncbi.nlm.nih.gov/39151981/

34. González R, Manun’Ebo MF, Meremikwu M, Rabeza VR, Sacoor C, Figueroa-Romero A, et al. The impact of community delivery of intermittent preventive treatment of malaria in pregnancy on its coverage in four sub-Saharan African countries (Democratic Republic of the Congo, Madagascar, Mozambique, and Nigeria): a quasi-experimental multicentre evaluation. Lancet Glob Health [Internet]. 2023 Apr 1 [cited 2025 Aug 3];11(4):e566–74. Available from: https://pubmed.ncbi.nlm.nih.gov/36925177/

35. WHO. World Health Organization. 2022 [cited 2025 Aug 3]. WHO Guidelines for malaria - 3 June 2022. Available from: http://apps.who.int/bookorders.

36. WHO. World Health Organization. 2023 [cited 2025 Aug 3]. Health Equity Assessment Toolkit. Available from: https://www.who.int/data/inequality-monitor/assessment_toolkit

37. Pons-Duran C, Llach M, Sacoor C, Sanz S, Macete E, Arikpo I, et al. Coverage of intermittent preventive treatment of malaria in pregnancy in four sub-Saharan countries: findings from household surveys. Int J Epidemiol [Internet]. 2020 Apr 1 [cited 2025 Aug 3];50(2):550. Available from: https://pmc.ncbi.nlm.nih.gov/articles/PMC8128463/

